# The Combination of Oxytocin with Mindfulness-Based Group Therapy Reduces Negative Symptoms in Schizophrenia Spectrum Disorders: A Triple-Blind, Placebo-Controlled, Randomized Clinical Pilot Trial (OXYMIND)

**DOI:** 10.64898/2026.07.01.26356996

**Authors:** Marco Zierhut, Max Alt, Inge Hahne, Niklas Bergmann, Felix Opper, Kristin Braun, Alice Braun, Julia Kraft, Thi Minh Tam Ta, Stephan Ripke, Malek Bajbouj, Eric Hahn, Kerem Böge

**Affiliations:** Charité – Universitätsmedizin Berlin, Corporate member of Freie Universität Berlin and Humboldt-Universität zu Berlin, Department of Psychiatry and Neuroscience, Berlin, Germany; Berlin Institute of Health at Charité – Universitätsmedizin Berlin, BIH Biomedical Innovation Academy, BIH Charité Clinician Scientist Program, Berlin, Germany; German Center for Mental Health (DZPG), partner site Berlin-Potsdam, Berlin-Potsdam, Germany; Stanley Centre for Psychiatric Research at Broad Institute of MIT and Harvard, Cambridge, MA, USA; Brandenburg Medical School, Department of Psychology, Neuruppin, Germany

**Keywords:** schizophrenia spectrum disorders, negative symptoms, oxytocin, mindfulness-based group therapy, clinical trial, positive symptoms

## Abstract

**Background:** Negative symptoms in schizophrenia spectrum disorders (SSD) remain insufficiently treated and require novel therapeutic approaches. Oxytocin may improve negative symptoms, although its effects appear highly context-dependent according to the social salience hypothesis. We conducted a randomized, triple-blind, placebo-controlled pilot study combining intranasal oxytocin with mindfulness-based group therapy (MBGT), hypothesizing that the positive social context of MBGT would enhance oxytocin-related effects.

**Methods:** 47 participants with SSD (34% female) were randomized to receive either 24 IU oxytocin (MBGT+OXT; n = 26) or placebo (MBGT+PLA; n = 21) before four MBGT sessions. Primary outcome was negative symptoms assessed with the Positive and Negative Syndrome Scale negative subscale (PANSS-N) at post-intervention and 4-week follow-up. Secondary outcomes included the Brief Negative Symptom Scale (BNSS), Self-Evaluation of Negative Symptoms Scale (SNS), and additional clinical measures. Linear mixed models estimated within- and between-group effects.

**Results:** Overall dropout rate was 14.89%, with one dropout potentially treatment-related. Blinding was successful. Participants completed 95.63% of sessions. Only the MBGT+OXT group showed significant improvements in PANSS-N from baseline to post-intervention (d = −0.74) and follow-up (d = −0.77), with a small between-group effect at follow-up (d = 0.39). BNSS total improved significantly only in the MBGT+OXT group from baseline to post-intervention (*d* = −0.88) and follow-up (*d* = −0.91), with between-group effects favoring MBGT+OXT at follow-up (d = −0.38). No serious adverse events occurred.

**Conclusions:** These findings suggest that oxytocin combined with MBGT may improve negative symptoms in SSD and support further large-scale trials.

**Clinical Trials Registration:** https://clinicaltrials.gov/study/NCT06136390, Registration number: NCT06136390

## Introduction

Schizophrenia spectrum disorders (SSDs) affect around 0.4% of the global population (1). and are characterized by positive, cognitive, affective symptoms, and negative symptoms, the latter remaining particularly difficult to treat (2). Negative symptoms are strongly associated with poor social functioning, reduced quality of life (3–5), and increased healthcare burden (6). More than half of individuals with SSDs present at least one prominent negative symptom (7), often emerging early (8) and persisting chronically (9), emphasizing the need for improved treatment strategies.

Current pharmacological and psychological interventions show limited efficacy for negative symptoms (10–12), prompting interest in pharmacologically augmented psychotherapies (13).

Specifically for negative symptoms, the hypothalamic neuropeptide oxytocin may emerge as promising augmentation strategy. While negative symptoms are associated with dysfunction in numerous neurobiological structures (14), recent models highlight networks for social reward anticipation and socioemotional processes (15, 16). In healthy individuals, oxytocin increases connection efficiency in these networks (17), is implicated in social cognition and empathy (18, 19), and is involved in social and non-social motivation related to avoidance and approach (20, 21).

Numerous studies have shown a dysregulated oxytocin system in SSD (22) and that improvements in social markers correlate with increases in oxytocin (23). Lower endogenous oxytocin levels are linked to more severe negative symptoms and impaired social cognition in SSD (24–27). The findings suggest oxytocin’s potential in SSD for negative symptoms. However, trials investigating oxytocin as an adjunct pharmacotherapy has yielded inconsistent results in improving social markers (28, 29), and negative symptoms (30). Most studies and meta-analyses have reported null effects (29, 31–35). According to the social salience hypothesis, oxytocińs effects are highly context dependent, suggesting that positive, supportive social contexts may be necessary to elicit therapeutic benefits (36). Five randomized controlled trials (RCTs) examining the augmentation of oxytocin in social skills or cognition training settings have failed to demonstrate additional benefits for negative symptoms beyond the training itself (37–41). These trainings were often intensive (up to 48 sessions) and may not have provided a definitively positive social context. To harness oxytocin’s potential in SSD, a more controllably positive, and non-directive setting may be necessary to elicit positive effects.

Targeting this need, mindfulness-based interventions (MBIs) are an established psychological approach, involving decentering attention and non-judgmental awareness of present sensations, thoughts, and emotions (42), recently being added to the German national treatment guidelines for schizophrenia (43). In SSD, mindfulness has been positively associated with positive emotions, anticipatory pleasure, and self-compassion (44, 45). Numerous meta-analyses have shown positive effects on positive and negative symptoms (46–48), with group settings being especially favourable (49) with participants experiencing mindfulness in a prosocial, interactive setting. Building on this evidence, our lab has pursued a stepwise translational research programme to adapt, implement, and refine mindfulness-based group therapy for people with SSD. In a first phase, the intervention was developed and manualized in a participatory manner, integrating clinical expertise, empirical evidence, and the perspectives of people with lived experience to ensure that mindfulness practices were acceptable, safe, and tailored to the specific needs of individuals with psychosis (50, 51). Our research lab has successfully trialled a twelve-session manualized inpatient mindfulness-based group therapy (MBGT) with significant, small effects on negative symptoms (52, 53) and changes in oxytocin levels (54). This inpatient work provided initial clinical and biological support for the intervention and informed the next translational step into less intensive outpatient settings, examining whether its social-affective mechanisms could be enhanced. We recently conducted a proof-of-concept feasibility RCT in an SSD outpatient setting and administered intranasal oxytocin before a two-session MBGT, compared with MBGT plus placebo (55). The combined intervention was feasible and exerted significant improvements, favouring the combination with oxytocin over placebo in self-reported negative affect and subjective stress. There was a trend (*p* = .06) and medium effect size (*η^2^_p_* = 0.09) for the combination of oxytocin with MGBT on the Self-Evaluation of Negative Symptoms (SNS) (56), with the *diminished emotional range* and *avolition* subscales reaching significance. Despite promising results, this trial was limited by the non-negative symptom primary outcome, lack of clinician-rated measures, and a short intervention period. Accordingly, the present study constitutes the next stage in this research pathway, extending prior participatory intervention development, inpatient implementation, and outpatient feasibility testing toward a longer, methodologically more rigorous, and mechanism-informed evaluation of oxytocin-augmented MBGT for negative symptoms in SSD.

Against this background, the present trial aimed to increase the combination effect by increasing the number of MBGT sessions to four, to enhance methodological robustness by adding a follow-up measurement to assess longer-term effects, and to employ clinician-rated measures of negative symptoms as primary outcome. Within a further triple-blinded pilot randomized controlled trial of this treatment approach, we examine the combined effect of oxytocin with MBGT for negative symptoms in outpatients with SSD.

## Methods and Materials

### Study design and Participants

This study used a triple-blind (participants, psychotherapists, raters), monocentric, randomized placebo-controlled pilot study design with four sessions of MBGT in combination with intranasal oxytocin (24 International Units [IU]) or placebo spray. The study was conducted at Charité – Universitätsmedizin Berlin, Campus Benjamin Franklin, in Germany between September 2023 and November 2024. It was approved by the corresponding ethics committee (EA4/196/19) and registered on clinicaltrials.gov (NCT06136390). Participants were recruited at the outpatient clinic of the Department of Psychiatry and Neurosciences (89%), externally from other psychiatric outpatient clinics, practices, and social services in Berlin, and through the clinic’s website (11%).

Inclusion criteria were 1) age 18-65, 2) diagnosis with a SSD (ICD-10: F2X.X), 3) proficiency in German language, and 4) informed consent. Exclusion criteria included 1) changes in psychiatric medication within two weeks before the first MBGT session, 2) a score > 5 on any item of the PANSS positive scale, 3) acute suicidality, 4) current substance use other than nicotine, 5) diagnosis of neurological disorders, 6) current pregnancy or breastfeeding, and 7) acute phase of electroconvulsive therapy.

### Procedures

As shown in Figure 1, study participation included baseline assessments within the week before treatment start, a four-week treatment period of weekly MBGT sessions, a post-intervention assessment (T1) within the week after the final MBGT session, and a follow-up assessment four weeks after treatment (T2).

**Figure 1:**
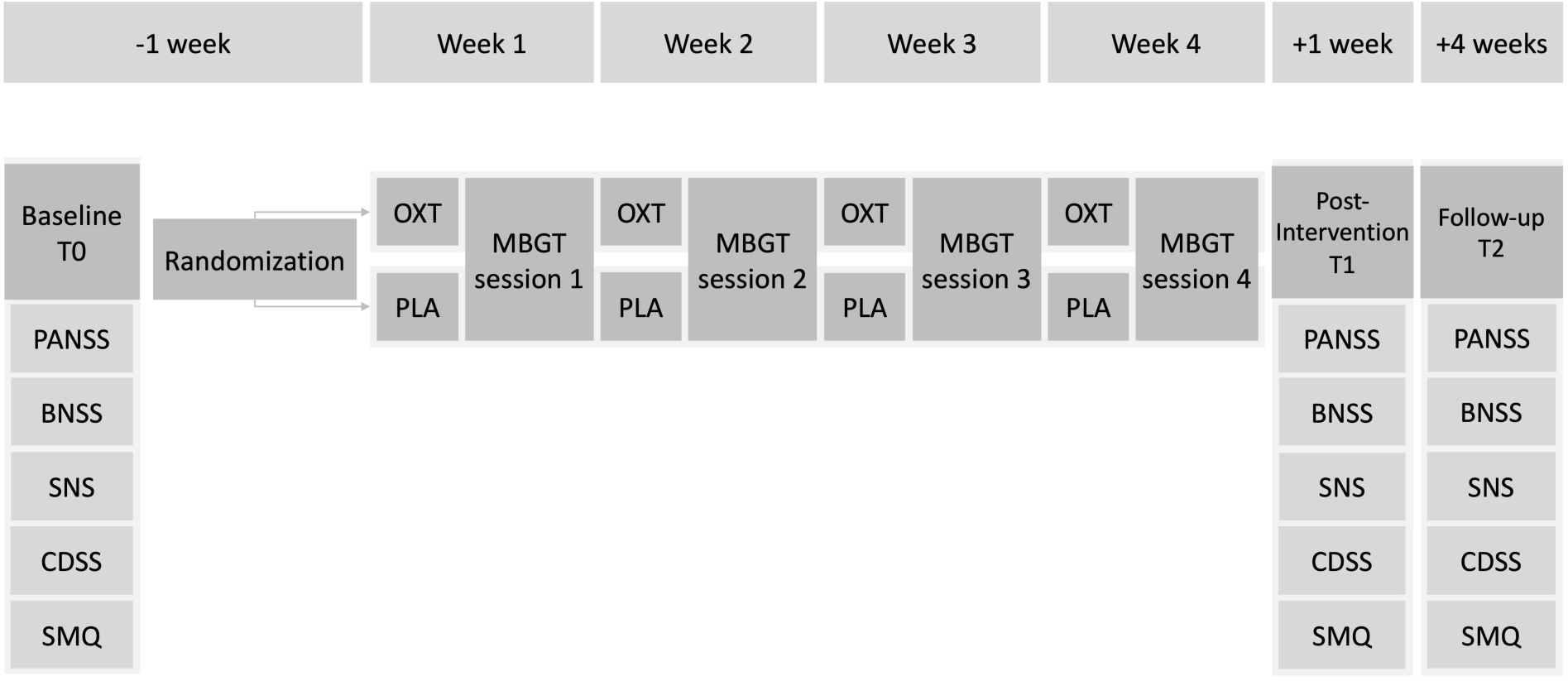
*Study design* Note. MBGT: Mindfulness-based group therapy; OXT: Oxytocin; PLA: Placebo; BNSS: Brief Negative Symptom Scale; PANSS-N: Positive and Negative Syndrome Scale - Negative Syndrome Scale; SNS: Self-Evaluation of Negative Symptoms.

Potentially eligible participants were contacted and screened for participation by telephone or in person. We requested participants bringing a medical report confirming their diagnosis.

Participants received written and verbal information about the study. After signing the consent form, sociodemographic information was collected. After baseline assessments, an independent researcher randomized all participants of the upcoming four sessions using a fixed-block size. Each participant in a coherent group setting received the same treatment. All participants continued to receive their usual treatments, although changes to medications were not permitted. Upon completion of the final assessment at follow-up, participants received €50 in compensation for their participation.

### Interventions

#### Oxytocin and Placebo

A research assistant administered six bursts of oxytocin or placebo nasal spray 30 minutes before each session. This interval aligns with plasma oxytocin pharmacokinetics, peaking 15–30 minutes after intranasal administration and returning to baseline by 60-90 minutes (57–59), adhering to the duration of our therapy. Participants in the experimental condition received synthetic oxytocin (24 IU Syntocinon®), whereas participants in the control group received a nasal spray containing saline. Spray bottles were not distinguishable by researchers or participants to ensure blinding.

### MBGT

Our manualized 12-session MBGT program (SENSE) was developed for a four-week treatment for inpatients with SSDs (53) and typically consists of 1 weekly 60-min session facilitated by a psychotherapist and two 30-minute sessions conducted by multidisciplinary interns. We adapted the manual to only include four 60-minute sessions, to ensure acceptability with outpatient participants (54), and to decrease time and stress in the context of applying oxytocin. Group sizes could vary from 3 to 6 participants. Each session covered one module of the manual, covering psychoeducational aspects and specific exercises, aiming to develop an understanding of key concepts of mindfulness and gain practical experience in a group. The modules included: 1) introduction to fundamentals of mindfulness and breathing exercises, 2) outdoor mindfulness and sensory perception, 3) releasing thoughts and non-judgement, and 4) mindfulness related to body awareness (60). Participants received non-mandatory homework, session-specific handouts, and were instructed to set small mindfulness-related goals for the upcoming week. Sessions were led by one MBGT-trained psychotherapist involved, with graduate-level psychology students assisting as co-therapists.

### Outcomes and Measures

Assessments included negative, positive and general symptoms, depressive symptoms, and mindfulness. Clinician-rated assessments were performed by seven trained and calibrated raters (medical doctors, medical doctorate students, psychology students). Figure 1 shows the assessment times throughout the trial. Baseline estradiol and prolactin levels were used in an outlier analysis due to interactions between oxytocin and sex hormones (61). Participants were recontacted after trial completion to enquire about their estimates of group allocation, to assess the success of participant blinding. Participants reporting that they were “unsure” were divided equally between the two groups.

### Negative Symptoms

The predefined primary outcomes of the study were within- and between-group changes of the clinician-rated German translation of the negative scale of the Positive and Negative Syndrome Scale (PANSS) from baseline to post-intervention and follow-up (62). Commonly used in clinical trials and practice, the PANSS has notorious shortcomings, including non-coverage of the five domains of the NIMH consensus on negative symptoms (63–65). Therefore, in accordance with European Psychiatric Association recommendations (66), we included the second-generation clinician-rated assessment Brief Negative Symptom Scale (BNSS) (67). Additionally, the second-generation self-rated measure Self-Evaluation of Negative Symptoms (SNS) was included (66). As supplemental analysis, we assessed PANSS Marder and Wallwork negative dimensions (68, 69).

### Positive, General, Depressive Symptoms, and Mindfulness

Further secondary clinical outcomes included PANSS total, positive, and general subscales, (70), clinician-rated depressive symptoms (Calgary Depression Scale for Schizophrenia: CDSS) (71), and self-rated mindfulness (Southampton Mindfulness Questionnaire; SMQ) (72, 73).

### Statistical methods

As a pilot study, we employed a pragmatic sample size estimation. Based on the previous between-group effect size (*η_p_^2^* = 0.09) on self-reported negative symptoms of our previous study (55), and recommendations for pilot studies, we attempted to recruit at least 15-25 individuals per group, anticipating small to medium effects (74). For within-group changes from baseline to post-treatment and baseline to follow-up, as well as within sessions, longitudinal linear mixed models were fitted. Fixed covariates included time (post-intervention and follow-up), group allocation, and group allocation*time interactions. Participants served as random intercepts. For between-group changes from baseline, longitudinal linear mixed-models were fitted to data including all participants with available measures post-baseline. Fixed covariates were fitted as above but additionally used baseline values as covariates. Differences within- or between-groups were examined using appropriate contrasts. Sample characteristics and outcome baseline scores were compared between groups using χ-squared tests for categorical- and independent samples *t*-tests for continuous variables to assess baseline imbalances. To assess the success of blinding, we calculated Cohen’s *κ,* with values between −0.2 and 0.2 (75).

As pilot study, we did not correct for multiple comparisons or perform imputations of missing values. We performed outlier analyses of serum prolactin and estradiol levels at baseline using Grubbs’ tests, stratified by sex (76). Serum oxytocin was not measured, as our previous study demonstrated a significant increase after administration (55).

Statistical analyses were performed in RStudio using the packages lme4, emmeans, and DeltaMAN (77–79), with a significance level of .05. Plots were created with the ggplot2 package (80). Pragmatic approximations of between-group Cohen’s d were calculated by dividing the mean differences by the pooled standard deviation at the relevant time-point. These were interpreted as small (*d* ≥ 0.2), medium (d ≥ 0.5), and large (d ≥ 0.8) (81).

## Results

### Sample description

The total sample comprised *N* = 47 participants, with *n* = 26 participants in the MBGT+OXT and *n* = 21 in the MBGT+PLA condition (see Figure 2). 13 cycles of four MBGT therapy sessions were conducted, seven in the MBGT+OXT and six in the MBGT+PLA conditions. Group sizes varied from 2-4 participants excluding dropouts: *M*_OXT_ = 3 (*SD* = 0.58); *M_PLA_* = 3.33 (*SD* = 1.03). The dropout rate at post-intervention was 19.2% (*n* = 5) in the MGBT+OXT and 9.52% (*n* = 2) in the MGBT+PLA condition, leading to an overall dropout rate of 14.89% over both groups. The study dropout rate remained the same at follow-up. Only one dropout in the MGBT+OXT condition was potentially treatment related. This participant reported a transient nasal irritation attributed to the administered nose spray. However, this did not fulfil the criteria for an adverse event. 38 participants (19 per group) were reached after trial completion. 32% and 55% of participants in the MBGT+OXT and MBGT+PLA groups correctly guessed their group, respectively. This led to a Cohen’s *κ* of −0.13 [95% CI: −0.44, 0.17], indicating successful blinding.

**Figure 2:**
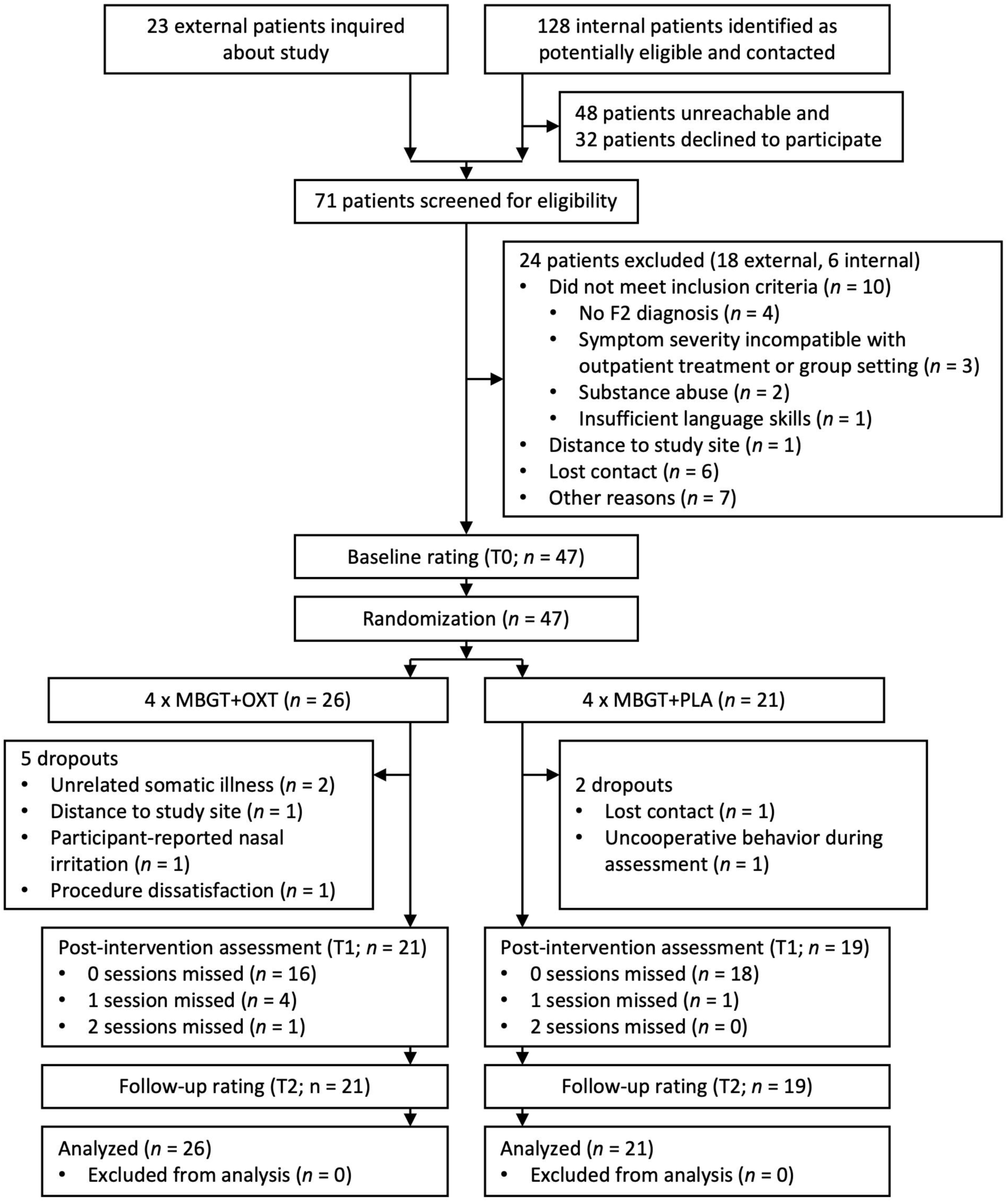
*Participant flow diagram* Note. MBGT: Mindfulness-based group therapy; OXT: Oxytocin; PLA: Placebo.

Retained participants completed 95.63% of all MBGT sessions, indicating strong acceptability of the treatment. Table 1 provides an overview of the sociodemographic and clinical characteristics at baseline. Baseline outcome measures were suitably balanced. However, the difference in rates of living with a partner (15% in MBGT+OXT vs. 33% in MBGT+PLA) was statistically significant. Considering this difference, a supplementary analysis was performed with this variable as a covariate (see the Supplement Table S2). The Grubbs’ tests did not reveal statistically significant outliers of prolactin and estradiol levels. Within-session stress and affect outcomes as well as group climate outcomes will be reported separately to focus on the primary outcomes in this context. Descriptive means and SDs for all outcomes can be found in the supplementary data Table S1.

**Table 1:**
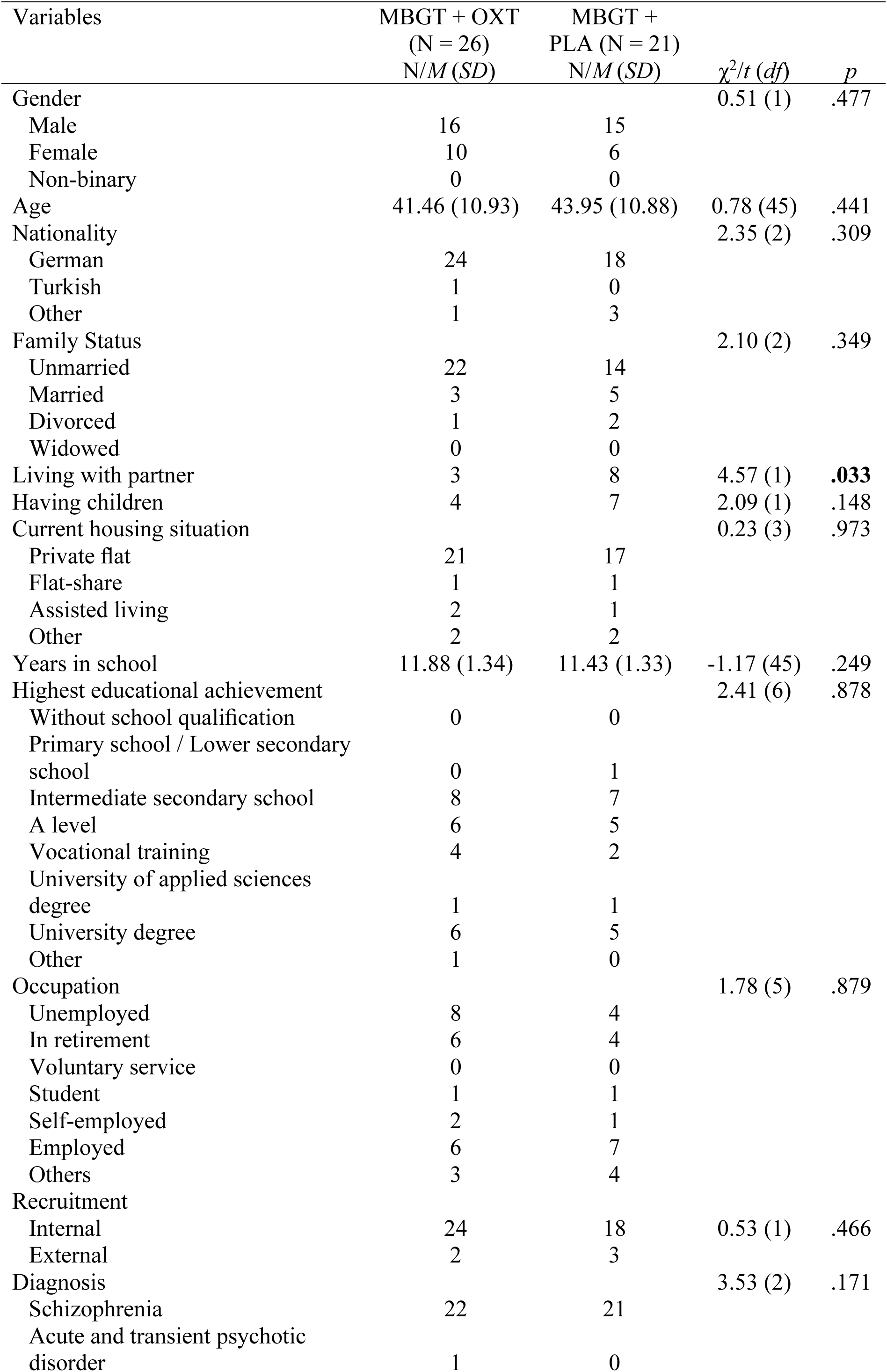
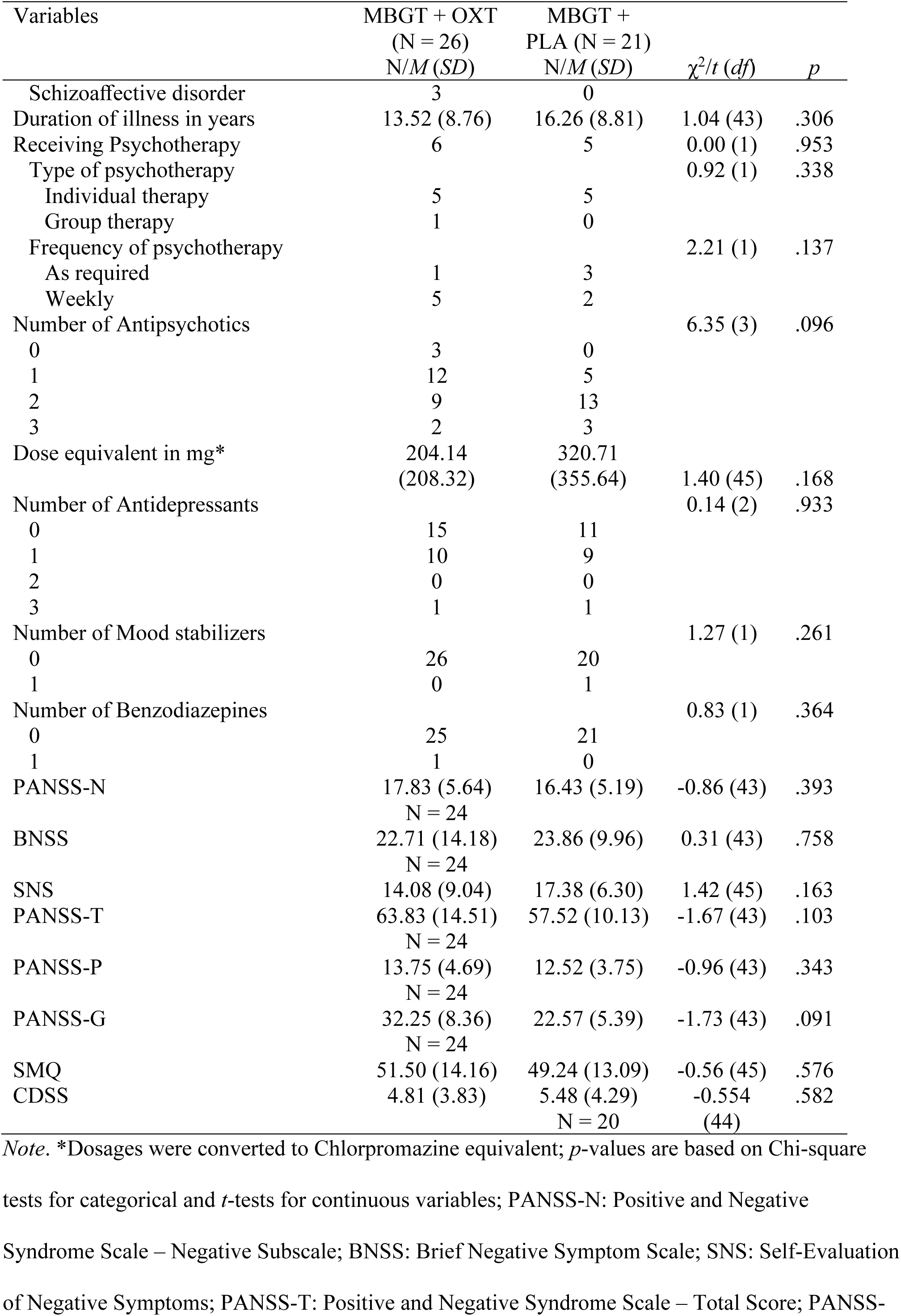
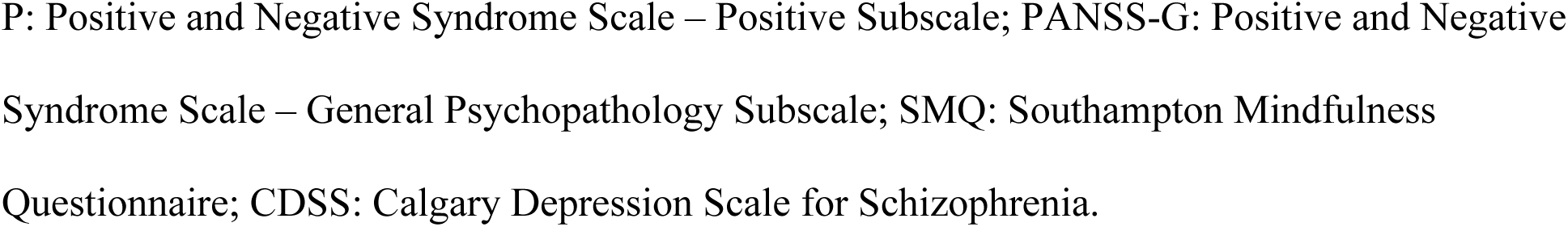
Sociodemographic, clinical data, and medication regime for both groups at baseline.

### Negative Symptoms

Within- and between-group differences for negative symptoms can be seen in Table 2 and 3, respectively, while Figure 3 shows plotted data. PANSS-N scores decreased significantly and with large effects sizes only in the MBGT+OXT group from baseline to post-intervention (Δ = −2.27, *p* < .001, *d* = −0.74) and baseline to follow-up (Δ = −2.75, *t* = −4.56, *p* <.001, *d =* −0.77). Concerning between-group differences, no significant differences were observed for PANSS-N at post-intervention (Δ = −1.73, *p* = .078, *d* = 0.30), but at follow-up, the model significantly favored MBGT+OXT (Δ = −2.08, *p* = .036) with a small effect size (*d* = 0.39), which can be explained by the small effects by the MBGT session itself in the placebo group. PANSS *Marder* and *Wallwork* negative scale dimensions showed similar effects.

**Figure 3:**
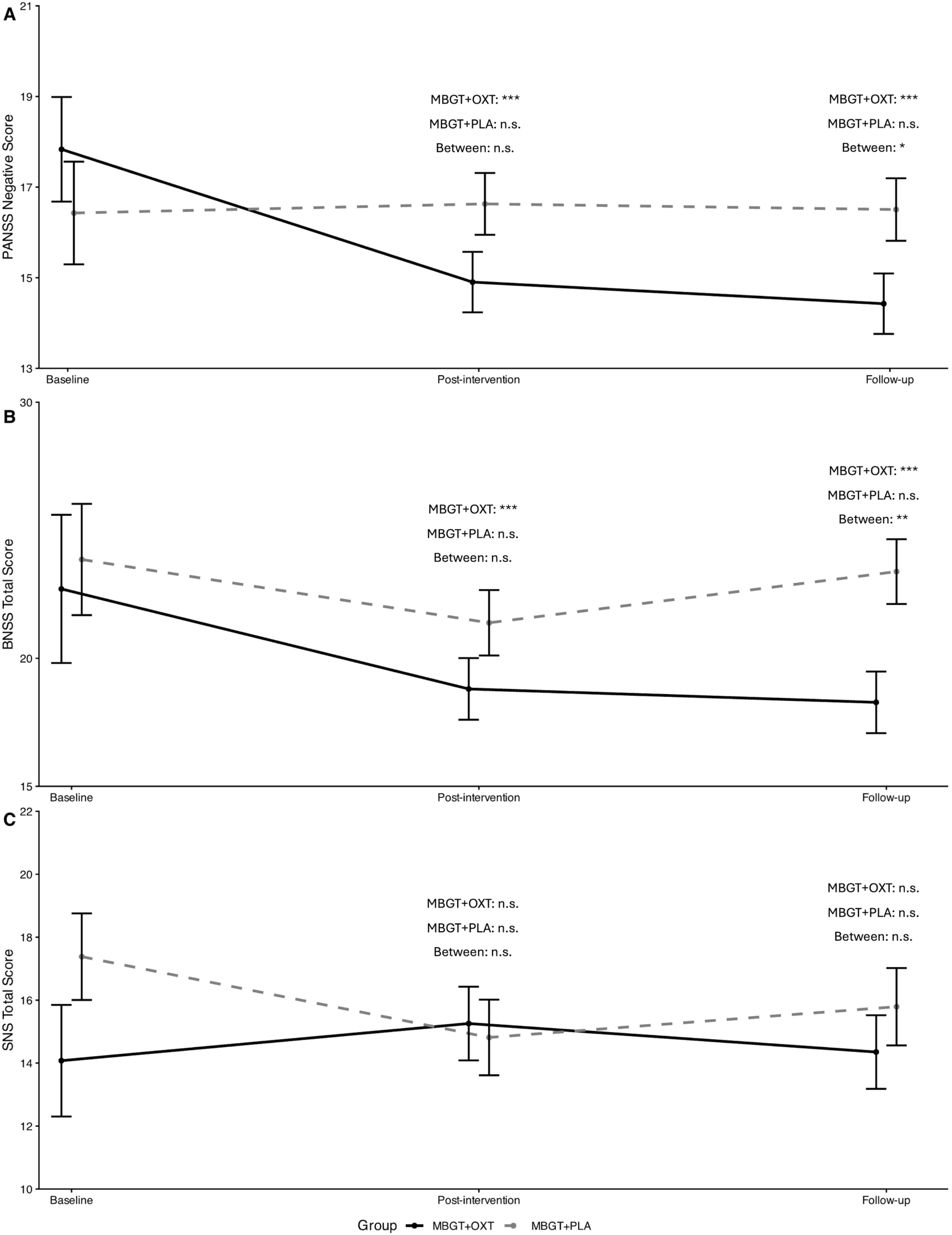
Plotted baseline and estimated marginal means with standard errors for PANSS-N (A), BNSS (B), and SNS (C) at post-intervention and follow-up in both groups Note. Between group: n.s. non-significant; * p < .05; ** p < .01; *** p < .001; PANSS-N: Positive and Negative Syndrome Scale - Negative Syndrome Scale; BNSS: Brief Negative Symptom Scale; SNS: Self-Evaluation of Negative Symptoms: T0: baseline; T1: post-intervention; T2: follow-up; MBGT: Mindfulness-based group therapy; OXT: Oxytocin; PLA: Placebo.

**Table 2:** Within-Group Changes across Baseline (T0), Post-Intervention (T1), and Four-Week Follow-Up (T2) for Negative Symptoms.

| **Group:**<br>**MBGT+** | **T1-T0** |  |  |  | **T2-T0** |  |  |  |
| --- | --- | --- | --- | --- | --- | --- | --- | --- |
|  | **Δ [95% CI]** | **t** | **p** | **d [95% CI]** | **Δ [95% CI]** | **t** | **p** | **d [95% CI]** |
| **PANSS-N** |  |  |  |  |  |  |  |  |
| OXT | -2.27 [-3.47, -1.07] | -3.77 | <.001 | -0.74 [-1.13, -0.35] | -2.75 [-3.95, -1.55] | -4.56 | <.001 | -0.77 [-1.1, -0.43] |
| PLA | -0.36 [-1.59, 0.87] | -0.58 | .562 | -0.12 [-0.51, 0.28] | -0.59 [-1.85, 0.66] | -0.94 | .349 | -0.25 [-0.79, 0.28] |
| **BNSS** |  |  |  |  |  |  |  |  |
| OXT | -4.53 [-6.66, -2.39] | -4.22 | <.001 | -0.88 [-1.3, -0.47] | -5.05 [-7.19, -2.91] | -4.71 | <.001 | -0.91 [-1.29, -0.52] |
| PLA | -2.12 [-4.41, 0.18] | -1.84 | .070 | -0.42 [-0.88, 0.04] | -0.03 [-2.26, 2.2] | -0.03 | .98 | 0 [-0.36, 0.35] |
| **BNSS Anhedonia** |  |  |  |  |  |  |  |  |
| OXT | -1.15 [-2.09, -0.21] | -2.44 | .017 | -0.6 [-1.1, -0.11] | -1.29 [-2.23, -0.36] | -2.74 | .008 | -0.63 [-1.09, -0.17] |
| PLA | -0.74 [-1.72, 0.25] | -1.48 | .142 | -0.29 [-0.68, 0.1] | 0.05 [-0.93, 1.04] | 0.11 | .914 | 0.02 [-0.32, 0.36] |
| **BNSS Distress** |  |  |  |  |  |  |  |  |
| OXT | 0.07 [-0.29, 0.43] | 0.38 | .702 | 0.09 [-0.39, 0.58] | -0.03 [-0.38, 0.33] | -0.15 | .884 | -0.03 [-0.44, 0.38] |
| PLA | -0.19 [-0.57, 0.18] | -1.02 | .309 | -0.19 [-0.55, 0.18] | -0.04 [-0.41, 0.34] | -0.19 | .851 | -0.04 [-0.45, 0.37] |
| **BNSS Asociality** |  |  |  |  |  |  |  |  |
| OXT | -0.56 [-1.14, 0.02] | -1.91 | .06 | -0.38 [-0.77, 0.02] | -0.6 [-1.18, -0.02] | -2.07 | .042 | -0.35 [-0.68, -0.01] |
| PLA | -0.23 [-0.84, 0.38] | -0.74 | .464 | -0.32 [-1.17, 0.54] | 0.25 [-0.36, 0.86] | 0.81 | .42 | 0.15 [-0.22, 0.53] |
| **BNSS Avolition** |  |  |  |  |  |  |  |  |
| OXT | -0.69 [-1.19, -0.2] | -2.8 | .006 | -0.56 [-0.96, -0.16] | -0.93 [-1.43, -0.44] | -3.76 | <.001 | -0.76 [-1.17, -0.36] |
| PLA | -0.58 [-1.09, -0.06] | -2.21 | .03 | -0.49 [-0.94, -0.05] | -0.21 [-0.73, 0.31] | -0.79 | .431 | -0.17 [-0.61, 0.26] |
| **BNSS Blunted affect** |  |  |  |  |  |  |  |  |
| OXT | -1.62 [-2.33, -0.9] | -4.5 | <.001 | -0.9 [-1.3, -0.5] | -1.66 [-2.38, -0.95] | -4.63 | <.001 | -0.94 [-1.35, -0.54] |
| PLA | -0.51 [-1.26, 0.25] | -1.34 | .183 | -0.43 [-1.08, 0.21] | -0.35 [-1.1, 0.4] | -0.93 | .358 | -0.17 [-0.55, 0.2] |
| **BNSS Alogia** |  |  |  |  |  |  |  |  |
| OXT | -0.56 [-1.18, 0.05] | -1.82 | .072 | -0.42 [-0.89, 0.04] | -0.52 [-1.13, 0.1] | -1.67 | .099 | -0.3 [-0.67, 0.06] |
| PLA | 0.12 [-0.54, 0.79] | 0.37 | .711 | 0.08 [-0.33, 0.48] | 0.24 [-0.41, 0.89] | 0.74 | .464 | 0.2 [-0.34, 0.74] |
| **SNS** |  |  |  |  |  |  |  |  |
| OXT | 0.38 [-2.21, 2.98] | 0.29 | .77 | 0.1 [-0.56, 0.75] | -0.52 [-3.12, 2.07] | -0.4 | .69 | -0.09 [-0.53, 0.35] |
| PLA | -1.12 [-3.81, 1.57] | -0.83 | .409 | -0.15 [-0.52, 0.22] | -0.14 [-2.88, 2.6] | -0.1 | .92 | -0.02 [-0.52, 0.47] |
| **SNS Social withdrawal** |  |  |  |  |  |  |  |  |
| OXT | 0.25 [-0.41, 0.91] | 0.76 | .451 | 0.18 [-0.29, 0.65] | 0.25 [-0.41, 0.91] | 0.76 | .451 | 0.17 [-0.27, 0.61] |
| PLA | -0.05 [-0.74, 0.63] | -0.16 | .874 | -0.03 [-0.41, 0.35] | -0.18 [-0.88, 0.51] | -0.53 | .597 | -0.12 [-0.56, 0.32] |
| **SNS Diminished emotional range** |  |  |  |  |  |  |  |  |
| OXT | 0.15 [-0.62, 0.92] | 0.39 | .701 | 0.1 [-0.4, 0.59] | 0.01 [-0.76, 0.78] | 0.02 | .986 | 0 [-0.34, 0.35] |
| PLA | -0.66 [-1.46, 0.14] | -1.63 | .106 | -0.41 [-0.9, 0.09] | 0 [-0.82, 0.82] | 0 | .998 | 0 [-0.6, 0.6] |
| **SNS Avolition** |  |  |  |  |  |  |  |  |
| OXT | -0.45 [-1.2, 0.3] | -1.19 | .238 | -0.29 [-0.78, 0.2] | -0.78 [-1.53, -0.03] | -2.07 | **.042** | -0.51 [-0.99, -0.02] |
| PLA | 0.01 [-0.77, 0.79] | 0.03 | .976 | 0.01 [-0.36, 0.38] | -0.15 [-0.94, 0.65] | -0.37 | .712 | -0.1 [-0.63, 0.43] |
| **SNS Anhedonia** |  |  |  |  |  |  |  |  |
| OXT | -0.08 [-0.86, 0.71] | -0.19 | .848 | -0.05 [-0.62, 0.51] | -0.22 [-1.01, 0.57] | -0.55 | .581 | -0.15 [-0.7, 0.4] |
| PLA | -0.17 [-0.99, 0.65] | -0.42 | .677 | -0.08 [-0.46, 0.3] | 0.08 [-0.75, 0.92] | 0.2 | .842 | 0.04 [-0.38, 0.47] |
| **SNS Alogia** |  |  |  |  |  |  |  |  |
| OXT | 0.47 [-0.33, 1.28] | 1.17 | .247 | 0.3 [-0.21, 0.8] | 0.19 [-0.62, 0.99] | 0.46 | .647 | 0.09 [-0.31, 0.49] |
| PLA | -0.27 [-1.11, 0.57] | -0.64 | .527 | -0.14 [-0.57, 0.3] | 0.08 [-0.78, 0.93] | 0.18 | .861 | 0.04 [-0.41, 0.49] |
| **PANSS-Marder** |  |  |  |  |  |  |  |  |
| OXT | -2.08 [-3.28, -0.87] | -3.43 | **.001** | -0.75 [-1.18, -0.31] | -2.46 [-3.67, -1.25] | -4.06 | **<.001** | -0.77 [-1.14, -0.39] |
| PLA | -0.92 [-2.16, 0.31] | -1.49 | .142 | -0.27 [-0.63, 0.09] | -0.73 [-1.99, 0.53] | -1.15 | .254 | -0.29 [-0.78, 0.21] |
| **PANSS-Wallwork** |  |  |  |  |  |  |  |  |
| OXT | -1.89 [-2.91, -0.87] | -3.68 | **<.001** | -0.84 [-1.3, -0.39] | -2.22 [-3.24, -1.2] | -4.33 | **<.001** | -0.8 [-1.16, -0.43] |
| PLA | -0.56 [-1.61, 0.49] | -1.07 | .289 | -0.19 [-0.55, 0.17] | -0.62 [-1.69, 0.45] | -1.15 | .252 | -0.31 [-0.85, 0.23] |

Note. MBGT: Mindfulness-based group therapy; OXT: Oxytocin; PLA: Placebo. PANSS-N: Positive and Negative Syndrome Scale – Negative
Subscale; BNSS: Brief Negative Symptom Scale; SNS: Self-Evaluation of Negative Symptoms; p-values below .05 are displayed in bold script, while those below .10 are displayed in italic script.

**Table 3:** Between-Group Changes across Baseline (T0), Post-Intervention (T1), and Four-Week Follow-Up (T2) for Negative Symptoms.

|  | T1 |  |  |  | T2 |  |  |  |
| --- | --- | --- | --- | --- | --- | --- | --- | --- |
| | $\Delta$ [95% CI] | t | p | d [95% CI] | $\Delta$ [95% CI] | t | p | d [95% CI] |
| **PANSS-N** | -1.73 [-3.66, 0.2] | -1.8 | .078 | -0.3 [-0.64, 0.04] | -2.08 [-4.02, -0.14] | -2.15 | **.036** | -0.39 [-0.76, -0.03] |
| **BNSS** | -2.58 [-6.12, 0.95] | -1.47 | .148 | -0.2 [-0.46, 0.07] | -5.11 [-8.61, -1.61] | -2.94 | **.005** | -0.38 [-0.64, -0.12] |
| **BNSS Anhedonia** | -0.52 [-1.98, 0.94] | -0.72 | .476 | -0.14 [-0.55, 0.26] | -1.45 [-2.91, 0] | -2.01 | .050 | -0.34 [-0.68, 0] |
| **BNSS Distress** | 0.25 [-0.26, 0.75] | 0.98 | .331 | 0.19 [-0.2, 0.58] | -0.01 [-0.51, 0.5] | -0.03 | .978 | 0 [-0.37, 0.36] |
| **BNSS Asociality** | -0.31 [-1.19, 0.57] | -0.71 | .478 | -0.13 [-0.51, 0.24] | -0.83 [-1.71, 0.04] | -1.91 | .062 | -0.32 [-0.66, 0.02] |
| **BNSS Avolition** | -0.12 [-0.89, 0.64] | -0.32 | .752 | -0.05 [-0.39, 0.28] | -0.73 [-1.49, 0.04] | -1.91 | .062 | -0.33 [-0.68, 0.02] |
| **BNSS Blunted affect** | -1.17 [-2.21, -0.12] | -2.24 | **.029** | -0.3 [-0.56, -0.03] | -1.37 [-2.42, -0.33] | -2.64 | **.011** | -0.38 [-0.66, -0.09] |
| **BNSS Alogia** | -0.7 [-1.64, 0.25] | -1.48 | .145 | -0.22 [-0.53, 0.08] | -0.73 [-1.67, 0.21] | -1.56 | .124 | -0.23 [-0.52, 0.06] |
| **SNS** | 0.44 [-2.94, 3.82] | 0.26 | .795 | 0.06 [-0.41, 0.54] | -1.44 [-4.85, 1.97] | -0.84 | .403 | -0.19 [-0.65, 0.27] |
| **SNS Social withdrawal** | 0.15 [-0.73, 1.03] | 0.34 | .733 | 0.09 [-0.45, 0.64] | 0.29 [-0.6, 1.18] | 0.65 | .515 | 0.16 [-0.32, 0.64] |
| **SNS Diminished emotional range** | 0.3 [-0.74, 1.33] | 0.57 | .568 | 0.18 [-0.44, 0.8] | -0.5 [-1.55, 0.54] | -0.96 | .341 | -0.24 [-0.74, 0.26] |
| **SNS Avolition** | -0.59 [-1.59, 0.41] | -1.17 | .245 | -0.25 [-0.69, 0.18] | -0.76 [-1.78, 0.25] | -1.5 | .138 | -0.34 [-0.78, 0.11] |
| **SNS Anhedonia** | 0.05 [-0.91, 1.02] | 0.11 | .91 | 0.03 [-0.45, 0.51] | -0.34 [-1.32, 0.64] | -0.69 | .491 | -0.15 [-0.6, 0.29] |
| **SNS Alogia** | 0.22 [-0.82, 1.27] | 0.43 | .671 | 0.12 [-0.44, 0.68] | -0.4 [-1.46, 0.65] | -0.76 | .449 | -0.21 [-0.75, 0.34] |
| **PANSS-Marder** | -0.98 [-2.8, 0.83] | -1.09 | .281 | -0.19 [-0.53, 0.16] | -1.61 [-3.44, 0.22] | -1.77 | *.083* | -0.28 [-0.61, 0.04] |
| **PANSS-Wallwork** | -1.18 [-2.69, 0.32] | -1.58 | .121 | -0.24 [-0.55, 0.07] | -1.47 [-2.99, 0.04] | -1.95 | *.057* | -0.3 [-0.61, 0.01] |

Note. MBGT: Mindfulness-based group therapy; OXT: Oxytocin; PLA: Placebo. PANSS-N: Positive and Negative Syndrome Scale – Negative Subscale; BNSS: Brief Negative Symptom Scale; SNS: Self-Evaluation of Negative Symptoms. Between-group differences below 0 represent a lower score in the MBGT+OXT group. p-values below .05 are displayed in bold script, while those below .10 are displayed in italic script.

Similar effects were seen for the BNSS total, which decreased significantly only in the MBGT+OXT group from baseline to post-intervention (Δ = −4.53, *p* < .001, *d* = −0.88) and baseline to follow-up (Δ = −5.05, *p* < .001, *d* = −0.91) with large effects sizes. Subscales *anhedonia, avolition,* and *blunted affect* improved significantly within the MBGT+OXT group from baseline to post-intervention and from baseline to follow-up, and *asociality* from baseline to follow-up. The subscale *avolition* improved for the MBGT+PLA group from baseline to post-intervention.

Concerning between-group effects of BNSS total, the model significantly favored MBGT+OXT at follow-up (Δ = −5.11, *p* = .005, *d* = −0.38), but not post-intervention (Δ = −2.58, *p* = .148, *d* = −0.2). Between-group effects were significant for the BNSS subscale *blunted affect* at post-intervention (Δ = −1.17, *p* = .029, *d* = −0.3) and follow-up (Δ = −1.37, *p* =.011, *d* = −0.38) with small effect sizes. For the subscale *anhedonia,* the between-group effect at follow-up bordered significance favoring MBGT+OXT (Δ = −1.45, *p* = .050, *d* = −0.34) with small effect size. The SNS total scale did not significantly decrease within either group, only the subscale *avolition* decreased within the MBGT-OXT group from baseline to follow-up with a medium effect size. No significant between-group effects were seen for the total SNS or its subscales at post-intervention (Δ = 0.44, *p* = .795, *d* = −0.06) or follow-up (Δ = −1.44, *p* = .403, *d* = 0.19).

### Positive, General, Depressive Symptoms, and Mindfulness

From baseline to post-intervention significant within-group changes were observed for the MBGT+OXT group for PANSS total (Δ = −6.44, *p* < .001, *d =* −0.84), general (Δ = −3.12, *p* = .002, *d* = −0.79) with large effect sizes and positive subscales (Δ = −1.06, *p* = .025, *d =* −0.5) with a medium effect size. From baseline to follow-up, MBGT+OXT showed significant improvements in the PANSS total (Δ = −8.06, *p* < .001, *d =* −0.99), general (Δ = −3.93, *p* < .001, *d* = −0.89), and positive (Δ = −1.39, *p =* .004, *d =* −0.82) with large effect sizes. Between-groups, PANSS total significantly improved at follow-up favoring MBGT+OXT with a small effect size (Δ = −5.54, *p* = .031, *d* = 0.45). No significant within- or between-group changes for depressive symptoms were found in the present study. The MBGT+OXT group showed a significant increase in the SMQ subscale *non-judgement* from baseline to post-intervention (Δ = 1.75, *p* = .029, *d* = 0.23) with small effect size. Within- and between-group results can be found in Tables 4 and 5.

**Table 4:**
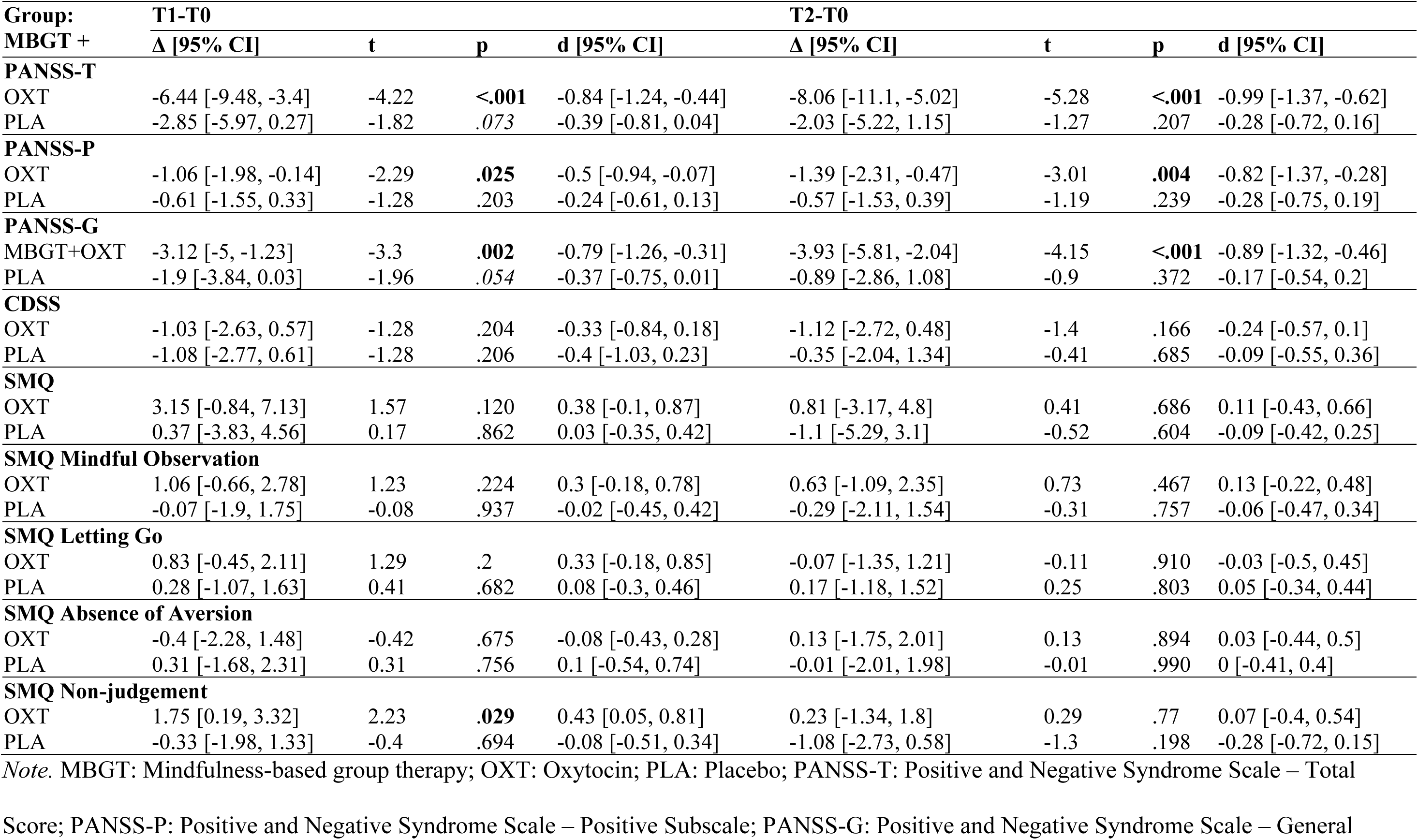
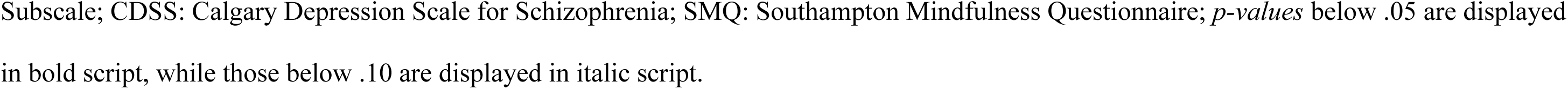
Within-Group Changes across Baseline (T0), Post-Intervention (T1), and Four-Week Follow-Up (T2) for Total Psychotic, Positive, General, Depressive Symptoms, and Mindfulness.

**Table 5:**
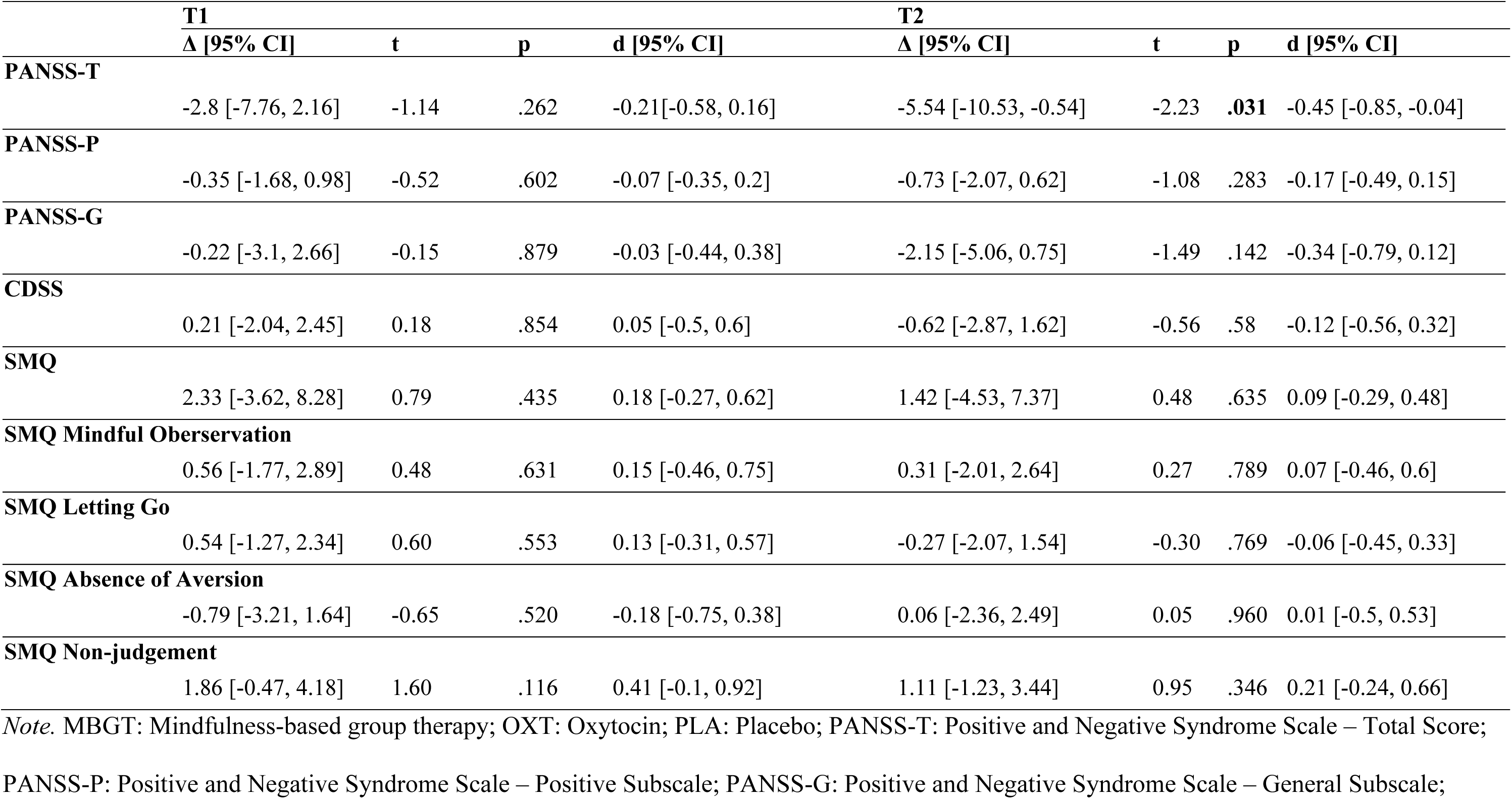
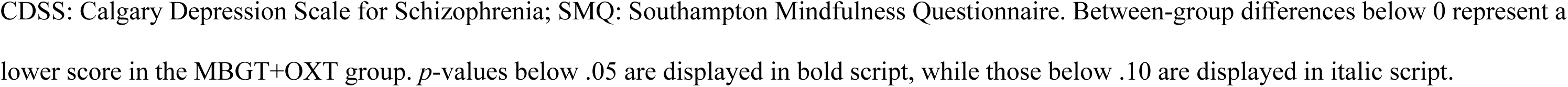
Between-Group Changes across Baseline (T0), Post-Intervention (T1), and Four-Week Follow-Up (T2) for Total Psychotic, Positive, General, Depressive Symptoms, and Mindfulness.

### Supplementary Analysis Correcting for Baseline Imbalance

As shown in the supplement Tables S2 and S3, controlling for baseline differences in *Living with a partner* did not decrease between-group effect sizes substantially for PANSS-N, but rendered the previously significant effect at follow-up non-significant. As effect sizes remained largely stable, this may be associated with the decreased degree of freedom. The significant between-group effect sizes at follow-up for BNSS-total and PANSS-total remained in place. The significant BNSS effect sizes of blunted affect at follow-up also remained significant.

## Discussion

The present randomized controlled triple-blind pilot trial compared a combination of a four-session mindfulness-based group therapy with 24 IU intranasal oxytocin (MBGT+OXT) to the combination with placebo (MBGT+PLA). Importantly, the intervention was safe, without serious adverse events attributable to treatment. Blinding was successful. Dropout rates were comparable to those reported for psychological interventions in SSD (14-20%) (82, 83), and acceptability was high (95.63%), indicating that participants experienced high satisfaction and commitment.

The pilot study design should be considered when interpreting the following results. Concerning our primary outcome of negative symptoms by the PANSS-N, the MBGT+OXT showed significant within-group changes from baseline to post-intervention and the follow-up, whereas MBGT+PLA did not. The PANSS negative symptom improvements in MBGT+OXT were also in the range of a clinically significant change (>1.5) (84), highlighting the clinical relevance of improvements. Furthermore, these improvements revealed a significant between-group effect size at the follow-up measurement, favoring MBGT+OXT. Consistent results were observed with the BNSS-total, with within-group changes exclusive to MBGT+OXT and a small but significant between-group effect at follow-up. Especially the subscale *blunted affect* demonstrated a significant between-group improvement favoring MBGT+OXT at post-intervention and follow-up, while *anhedonia* approached significance at follow-up. These two subscales may be the strongest drivers of negative symptom improvements.

As no improvements were seen in the clinician-rated depression scale CDSS, the improvements may encouragingly point to direct and difficult to achieve improvements in primary negative symptoms as opposed to secondary negative symptoms occurring in the context of depressive symptoms (71). Further research may more conclusively delineate the mechanistic relevance of oxytocin to these symptoms. It should also be noted that the blinding in this combination study was successful, strengthening the interpretation of the results facing the frequently criticised unblinding bias in augmentation studies.

Improvements in positive, total, and general symptoms were also observed exclusively in the MBGT+OXT group, with between-group significance at follow-up for total symptoms with a small effect size. Previous studies and meta-research have found improvements in these symptoms with mindfulness-based approaches on their own (48), whereas improvements have been inconsistent with sole oxytocin administration (33). These findings highlight the potential of oxytocin and MBGT beyond negative symptoms, warranting further investigation. It should be noted that this study involved only a four-week course of treatment comprising four sessions. It may therefore be particularly relevant to explore the potential for extending the treatment period to eight, twelve or more weeks.

Between-group differences in PANSS-N and BNSS became significant at follow-up measurements, potentially due to oxytocin’s effect in increasing social cognitive connectivity in cortex regions associated with social reward (85), in interplay with the positive setting and effects of the MBGT-sessions. It is not surprising that there were no significant changes in negative symptoms in the MBGT+PLA group, as effects of MBGT have generally been observed in studies comprising eight sessions, whereas our pilot outpatient study involved only four sessions (52). Additionally, relevant mechanisms of mindfulness-based interventions for negative symptoms include increasing psychological flexibility (86), improved stress regulation and stress biomarkers like cortisol (87), heightened positive affect and anticipatory pleasure (49), and increased interest and engagement in social activities (88). Oxytocin’s role in enhancing social functioning in a context-dependent manner may have amplified and prolonged these effects (36). Further investigation into variables such as group climate, stress, affect, social functioning, and mindfulness as possible mediators of MBGT and oxytocin on negative and symptoms is warranted for clearer evidence concerning mechanisms of change.

Our findings contrast with null findings of previous group therapies augmented with oxytocin for negative symptoms (37–41). The manuals employed in other trials were often highly structured skills-training interventions (89). In contrast, MBGT cultivates emotional awareness, non-judgment, and interpersonal safety through experiential learning (53). This may have created a safe and pleasant group environment necessary for oxytocin to elicit its prosocial effects. The results partially replicate those of our previous pilot study (55), which showed significant between-group effects for self-rated negative symptoms using SNS, particularly in the *diminished emotional range* and *avolition subscales*. In the current trial, no between-group effects were observed for self-rated negative symptoms on the SNS, in contrast to the results for the clinician-rated measures.

Only the subscale *avolition* improved within the MBGT+OXT group from baseline to follow-up. The previous pilot study employed a different study design, including a shorter intervention period of only two sessions in one week and no follow-up, which may have contributed to this difference. Other factors, such as insight into negative symptoms (90) or cognitive functioning (91), may also be associated with the non-convergence between self-reported and clinician-rated negative symptoms. Future research may offer clearer insights regarding comparisons of self-report and clinician-rated measures of negative symptoms.

This pilot study’s promising results justify sufficiently powered, large-scale, multicenter RCTs of MBGT combined with oxytocin, incorporating longer follow-up periods. Consistent findings from further trials could inform the implementation of this approach in SSD as novel, safe, and effective treatment for hard-to-treat chronic negative symptoms.

## Strengths and limitations

The results of this trial should be considered alongside its limitations. Firstly, the pilot study sample size was small, likely resulting in underpowered analyses. Therefore, it should be interpreted with caution, given corrections for multiple testing were not applied. Future studies should be designed adequately powered. Secondly, the monocentric and exclusively outpatient-oriented design limits the external validity of the findings. Lastly, oxytocin was limited to 24 IU compared with placebo. The dosage should be varied in future studies to investigate dosage effects. Some previous studies found oxytocin’s effects to be more apparent between 36 and 48 IU for social processing (92), and above 40 IU for negative symptoms (35), although positive results for higher-order social functions have been shown at 24 IU (93).

Nevertheless, multiple strengths are apparent: The triple-blinded design (participants, psychotherapists, raters) reduced bias. The MBGT treatment is based on a standardized intervention with high feasibility and acceptability, adapted to the outpatient setting. Negative symptoms were assessed using multiple instruments, in accordance with recommendations for their assessment (66). Blinding was successful, which further highlights reduced bias in outcomes. Lastly, we included female participants, robustly checking for pregnancy, breastfeeding, and outliers in the sex hormones prolactin and estradiol, which may affect oxytocin effects (61). Many other studies investigating the effects of exogenous oxytocin omit including female participants or controlling for sex hormone outliers (94).

## Conclusion

This triple-blinded randomized controlled pilot trial successfully implemented oxytocin combined with MBGT as a promising approach for improving hard-to-treat, often chronic negative symptoms in individuals with SSD. Our findings suggest that the combination of oxytocin with MBGT as positive social context may have the potential to reduce negative symptoms. Given the pilot nature of this study, rigorous large-scale RCTs are required to replicate and further contextualise our findings.

## Data availability

The data and code that support the findings of this study are available from the corresponding author upon reasonable request.

## Acknowledgements

Dr. Marco Zierhut is a participant in the Berlin Institute of Health (BIH) Charité Clinician Scientist Program funded by the Charité – Universitätsmedizin Berlin, and the Berlin Institute of Health at Charité (BIH), Germany. This work is dedicated to Dr Renaldo Bernard, who, as a researcher and friend, made a significant contribution to this manuscript through his constant support, and whose contribution is to be honoured in this way following his unexpected death.

