## Supplement for "The Combination of Oxytocin with Mindfulness-Based Group Therapy Reduces Negative Symptoms in Schizophrenia Spectrum Disorders: A Triple-Blind, Placebo-Controlled, Randomized Clinical Pilot Trial (OXYMIND)"

**Table S1** – *Descriptive Means, Standard Deviations, and Sample Sizes for Outcomes Measured at Baseline, Post-Intervention, and Follow-up*

| | | Baseline (T0) | | Post-Intervention (T1) | | 4-Week Follow-up (T2) | |
|---|---|---|---|---|---|---|---|
| **Outcome** | **Group** | **Mean (SD)** | **N** | **Mean (SD)** | **N** | **Mean (SD)** | **N** |
| **PANSS-N** | | | | | | | |
| | OXT | 17.83 (5.65) | 24 | 15.67 (5.18) | 21 | 15.19 (5.36) | 21 |
| | PLA | 16.43 (5.19) | 21 | 16.00 (6.20) | 20 | 15.21 (5.22) | 19 |
| **BNSS** | | | | | | | |
| | OXT | 22.71 (14.18) | 24 | 18.71 (14.03) | 21 | 18.19 (13.71) | 21 |
| | PLA | 23.86 (9.96) | 21 | 21.28 (12.14) | 18 | 23.47 (13.05) | 19 |
| **BNSS Anhedonia** | | | | | | | |
| | OXT | 5.96 (4.21) | 24 | 4.76 (3.66) | 21 | 4.62 (4.12) | 21 |
| | PLA | 6.62 (3.80) | 21 | 5.58 (3.52) | 19 | 6.37 (4.40) | 19 |
| **BNSS Distress** | | | | | | | |
| | OXT | 0.79 (1.38) | 24 | 0.95 (1.47) | 21 | 0.86 (1.53) | 21 |
| | PLA | 0.86 (1.49) | 21 | 0.74 (1.05) | 19 | 0.89 (1.20) | 19 |
| **BNSS Asociality** | | | | | | | |
| | OXT | 3.08 (2.36) | 24 | 2.62 (2.11) | 21 | 2.57 (2.23) | 21 |
| | PLA | 3.05 (2.69) | 21 | 2.74 (2.54) | 19 | 3.21 (2.92) | 19 |
| **BNSS Avolition** | | | | | | | |
| | OXT | 3.62 (2.20) | 24 | 2.76 (2.43) | 21 | 2.52 (2.16) | 21 |
| | PLA | 3.76 (1.84) | 21 | 3.21 (2.10) | 19 | 3.58 (2.24) | 19 |
| **BNSS Blunted affect** | | | | | | | |
| | OXT | 6.33 (4.08) | 24 | 5.19 (3.93) | 21 | 5.14 (3.57) | 21 |
| | PLA | 6.86 (3.71) | 21 | 6.53 (3.98) | 19 | 6.68 (3.71) | 19 |
| **BNSS Alogia** | | | | | | | |
| | OXT | 2.92 (3.61) | 24 | 2.43 (3.41) | 21 | 2.48 (3.23) | 21 |
| | PLA | 2.71 (2.74) | 21 | 2.39 (2.75) | 18 | 2.74 (3.16) | 19 |
| **SNS** | | | | | | | |
| | OXT | 14.08 (9.04) | 26 | 14.14 (7.60) | 21 | 13.24 (8.15) | 21 |
| | PLA | 17.38 (6.30) | 21 | 16.10 (6.49) | 20 | 16.89 (6.50) | 19 |
| **SNS Social withdrawal** | | | | | | | |
| | OXT | 2.58 (1.86) | 26 | 2.76 (1.73) | 21 | 2.76 (1.84) | 21 |
| | PLA | 2.95 (1.88) | 21 | 2.80 (1.47) | 20 | 2.58 (1.87) | 19 |

| | | | | | | | |
|---|---|---|---|---|---|---|---|
| **SNS Diminished emotional range** | | | | | | | |
| | OXT | 2.62 (2.02) | 26 | 2.71 (1.82) | 21 | 2.57 (2.27) | 21 |
| | PLA | 3.86 (1.77) | 21 | 3.25 (1.48) | 20 | 3.89 (1.85) | 19 |
| **SNS Avolition** | | | | | | | |
| | OXT | 3.31 (2.46) | 26 | 2.81 (2.48) | 21 | 2.48 (2.44) | 21 |
| | PLA | 3.81 (2.02) | 21 | 3.85 (2.13) | 20 | 3.68 (2.06) | 19 |
| **SNS Anhedonia** | | | | | | | |
| | OXT | 3.65 (2.45) | 26 | 3.52 (1.94) | 21 | 3.38 (2.20) | 21 |
| | PLA | 3.76 (2.17) | 21 | 3.55 (2.06) | 20 | 3.79 (2.20) | 19 |
| **SNS Alogia** | | | | | | | |
| | OXT | 1.92 (2.19) | 26 | 2.33 (1.98) | 21 | 2.05 (1.99) | 21 |
| | PLA | 3.00 (1.87) | 21 | 2.65 (1.73) | 20 | 2.95 (1.90) | 19 |
| **PANSS-T** | | | | | | | |
| | OXT | 63.83 (14.51) | 24 | 57.33 (14.66) | 21 | 55.71 (14.57) | 21 |
| | PLA | 57.52 (10.14) | 21 | 54.10 (11.98) | 20 | 53.58 (9.43) | 19 |
| **PANSS-P** | | | | | | | |
| | OXT | 13.75 (4.69) | 24 | 12.62 (5.63) | 21 | 12.29 (4.91) | 21 |
| | PLA | 12.52 (3.75) | 21 | 11.85 (3.80) | 20 | 11.47 (3.31) | 19 |
| **PANSS-G** | | | | | | | |
| | OXT | 32.25 (8.36) | 24 | 29.05 (8.29) | 21 | 28.24 (7.71) | 21 |
| | PLA | 28.57 (5.39) | 21 | 26.25 (5.44) | 20 | 26.89 (4.45) | 19 |
| **CDSS** | | | | | | | |
| | OXT | 5.48 (4.29) | 25 | 4.57 (4.58) | 21 | 4.48 (5.51) | 21 |
| | PLA | 4.81 (3.83) | 21 | 3.58 (3.49) | 19 | 4.32 (4.64) | 19 |
| **SMQ** | | | | | | | |
| | OXT | 44.50 (14.16) | 26 | 48.00 (15.88) | 21 | 45.67 (17.12) | 21 |
| | PLA | 46.76 (13.09) | 21 | 45.74 (9.69) | 19 | 45.00 (13.25) | 19 |
| **SMQ Mindful Observation** | | | | | | | |
| | OXT | 10.58 (4.68) | 26 | 11.86 (4.87) | 21 | 11.43 (4.91) | 21 |
| | PLA | 11.71 (3.44) | 21 | 11.47 (2.20) | 19 | 11.21 (3.66) | 19 |
| **SMQ Letting Go** | | | | | | | |
| | OXT | 12.42 (4.47) | 26 | 13.48 (4.75) | 21 | 12.57 (5.21) | 21 |
| | PLA | 12.57 (4.14) | 21 | 12.53 (3.17) | 19 | 12.58 (3.82) | 19 |
| **SMQ Absence of Aversion** | | | | | | | |
| | OXT | 10.08 (4.80) | 26 | 9.71 (4.50) | 21 | 10.24 (4.36) | 21 |

| | | | | | | | |
|---|---|---|---|---|---|---|---|
| | PLA | 10.24 (4.27) | 21 | 10.42 (4.09) | 19 | 10.26 (5.09) | 19 |
| **SMQ Non-judgement** | | | | | | | |
| | OXT | 11.42 (4.25) | 26 | 12.95 (5.59) | 21 | 11.43 (5.83) | 21 |
| | PLA | 12.24 (4.23) | 21 | 11.32 (2.93) | 19 | 10.95 (4.42) | 19 |
| **PANSS-Marder** | | | | | | | |
| | OXT | 17.96 (5.86) | 24 | 16.00 (5.39) | 21 | 15.62 (5.67) | 21 |
| | PLA | 17.10 (5.54) | 21 | 16.00 (5.23) | 20 | 15.95 (5.64) | 19 |
| **PANSS-Wallwork** | | | | | | | |
| | OXT | 15.75 (5.24) | 24 | 14.00 (4.73) | 21 | 13.67 (4.89) | 21 |
| | PLA | 14.95 (4.85) | 21 | 14.30 (5.01) | 20 | 14.00 (4.92) | 19 |

*Note.* MBGT: Mindfulness-based group therapy; OXT: Oxytocin; PLA: Placebo; PANSS-N: Positive and Negative Syndrome Scale – Negative Subscale; BNSS: Brief Negative Symptom Scale; SNS: Self-Evaluation of Negative Symptoms; PANSS-T: Positive and Negative Syndrome Scale – Total Score; PANSS-P: Positive and Negative Syndrome Scale – Positive Subscale; PANSS-G: Positive and Negative Syndrome Scale – General Psychopathology Subscale; CDSS: Calgary Depression Scale for Schizophrenia; SMQ: Southampton Mindfulness Questionnaire.

**Table S2** – *Within Group Changes across Baseline (T0), Post-Intervention (T1), and Four-Week Follow-Up (T2) including "living with partner" as covariate*

| Outcome/Group | Within Group Effects | | | | | | | |
|---|---|---|---|---|---|---|---|---|
| | **T1-T0** | | | | **T2-T0** | | | |
| | **Δ [95% CI]** | **t** | **p** | **d [95% CI]** | **Δ [95% CI]** | **t** | **p** | **d [95% CI]** |
| **PANSS-N** | | | | | | | | |
| OXT | -2.27 [-3.47, -1.07] | -3.77 | **<.001** | -0.74 [-1.13, -0.35] | -2.75 [-3.95, -1.55] | -4.56 | **<.001** | -0.77 [-1.1, -0.43] |
| PLA | -0.36 [-1.59, 0.87] | -0.58 | .561 | -0.12 [-0.51, 0.28] | -0.59 [-1.85, 0.66] | -0.94 | .35 | -0.25 [-0.79, 0.28] |
| **BNSS** | | | | | | | | |
| OXT | -4.53 [-6.66, -2.39] | -4.22 | **<.001** | -0.88 [-1.3, -0.47] | -5.05 [-7.19, -2.91] | -4.71 | **<.001** | -0.91 [-1.29, -0.52] |
| PLA | -2.11 [-4.41, 0.18] | -1.84 | *.07* | -0.42 [-0.88, 0.04] | -0.03 [-2.28, 2.22] | -0.02 | .981 | 0 [-0.36, 0.36] |
| **BNSS Anhedonia** | | | | | | | | |
| OXT | -1.15 [-2.09, -0.21] | -2.44 | **.017** | -0.6 [-1.1, -0.11] | -1.29 [-2.23, -0.35] | -2.74 | .008 | -0.63 [-1.09, -0.17] |
| PLA | -0.73 [-1.72, 0.25] | -1.48 | .143 | -0.29 [-0.68, 0.1] | 0.06 [-0.93, 1.04] | 0.11 | .912 | 0.02 [-0.32, 0.36] |
| **BNSS Distress** | | | | | | | | |
| OXT | 0.07 [-0.29, 0.43] | 0.38 | .708 | 0.09 [-0.39, 0.57] | -0.03 [-0.39, 0.33] | -0.15 | .878 | -0.03 [-0.45, 0.38] |
| PLA | -0.19 [-0.57, 0.18] | -1.03 | .307 | -0.19 [-0.55, 0.18] | -0.04 [-0.41, 0.34] | -0.19 | .848 | -0.04 [-0.45, 0.37] |
| **BNSS Asociality** | | | | | | | | |
| OXT | -0.56 [-1.14, 0.02] | -1.91 | *.06* | -0.38 [-0.77, 0.02] | -0.6 [-1.18, -0.02] | -2.08 | **.041** | -0.35 [-0.68, -0.01] |
| PLA | -0.23 [-0.84, 0.38] | -0.74 | .464 | -0.32 [-1.17, 0.54] | 0.25 [-0.36, 0.86] | 0.81 | .42 | 0.15 [-0.22, 0.53] |
| **BNSS Avolition** | | | | | | | | |
| OXT | -0.69 [-1.18, -0.2] | -2.79 | **.007** | -0.56 [-0.96, -0.16] | -0.93 [-1.42, -0.44] | -3.75 | **<.001** | -0.76 [-1.17, -0.36] |
| PLA | -0.57 [-1.09, -0.05] | -2.2 | **.031** | -0.49 [-0.93, -0.05] | -0.21 [-0.72, 0.31] | -0.79 | .434 | -0.17 [-0.61, 0.27] |
| **BNSS Blunted affect** | | | | | | | | |
| OXT | -1.61 [-2.33, -0.9] | -4.49 | **<.001** | -0.9 [-1.3, -0.5] | -1.66 [-2.38, -0.95] | -4.62 | **<.001** | -0.94 [-1.35, -0.54] |
| PLA | -0.51 [-1.26, 0.25] | -1.34 | .185 | -0.43 [-1.08, 0.21] | -0.35 [-1.1, 0.4] | -0.92 | .359 | -0.17 [-0.55, 0.2] |
| **BNSS Alogia** | | | | | | | | |
| OXT | -0.57 [-1.18, 0.05] | -1.83 | *.072* | -0.43 [-0.89, 0.04] | -0.52 [-1.13, 0.1] | -1.67 | *.098* | -0.31 [-0.67, 0.06] |
| PLA | 0.12 [-0.54, 0.78] | 0.37 | .716 | 0.07 [-0.33, 0.48] | 0.24 [-0.41, 0.89] | 0.73 | .465 | 0.2 [-0.34, 0.74] |
| **SNS** | | | | | | | | |
| OXT | 0.37 [-2.23, 2.96] | 0.28 | .78 | 0.09 [-0.56, 0.74] | -0.54 [-3.13, 2.06] | -0.41 | .68 | -0.09 [-0.54, 0.35] |
| PLA | -1.15 [-3.84, 1.54] | -0.85 | .398 | -0.16 [-0.53, 0.21] | -0.16 [-2.89, 2.58] | -0.11 | .91 | -0.03 [-0.52, 0.46] |
| **SNS Social withdrawal** | | | | | | | | |
| OXT | 0.25 [-0.41, 0.9] | 0.74 | .459 | 0.18 [-0.3, 0.65] | 0.25 [-0.41, 0.9] | 0.74 | .459 | 0.16 [-0.28, 0.61] |

| | | | | | | | | |
|---|---|---|---|---|---|---|---|---|
| PLA | -0.06 [-0.74, 0.62] | -0.18 | .858 | -0.03 [-0.41, 0.34] | -0.19 [-0.88, 0.5] | -0.54 | .588 | -0.12 [-0.56, 0.32] |
| **SNS Diminished emotional range** | | | | | | | | |
| OXT | 0.15 [-0.62, 0.92] | 0.39 | .7 | 0.1 [-0.4, 0.59] | 0.01 [-0.76, 0.78] | 0.02 | .986 | 0 [-0.34, 0.35] |
| PLA | -0.66 [-1.46, 0.14] | -1.64 | .106 | -0.41 [-0.9, 0.09] | 0 [-0.82, 0.82] | <.001 | .999 | 0 [-0.6, 0.6] |
| **SNS Avolition** | | | | | | | | |
| OXT | -0.45 [-1.2, 0.3] | -1.2 | .234 | -0.3 [-0.78, 0.19] | -0.79 [-1.54, -0.04] | -2.08 | **.04** | -0.51 [-0.99, -0.02] |
| PLA | 0.01 [-0.77, 0.78] | 0.01 | .99 | 0 [-0.37, 0.37] | -0.15 [-0.94, 0.64] | -0.38 | .704 | -0.1 [-0.63, 0.43] |
| **SNS Anhedonia** | | | | | | | | |
| OXT | -0.08 [-0.87, 0.71] | -0.21 | .836 | -0.06 [-0.62, 0.51] | -0.23 [-1.01, 0.56] | -0.57 | .571 | -0.16 [-0.7, 0.39] |
| PLA | -0.18 [-1, 0.64] | -0.44 | .661 | -0.08 [-0.47, 0.3] | 0.08 [-0.75, 0.91] | 0.19 | .853 | 0.04 [-0.38, 0.46] |
| **SNS Alogia** | | | | | | | | |
| OXT | 0.47 [-0.34, 1.27] | 1.15 | .252 | 0.29 [-0.21, 0.8] | 0.18 [-0.62, 0.99] | 0.45 | .655 | 0.09 [-0.31, 0.49] |
| PLA | -0.28 [-1.12, 0.57] | -0.65 | .516 | -0.14 [-0.58, 0.29] | 0.07 [-0.78, 0.93] | 0.17 | .868 | 0.04 [-0.41, 0.49] |
| **PANSS-T** | | | | | | | | |
| OXT | -6.45 [-9.49, -3.41] | -4.22 | **<.001** | -0.84 [-1.24, -0.45] | -8.07 [-11.11, -5.03] | -5.28 | **<.001** | -0.99 [-1.37, -0.62] |
| PLA | -2.86 [-5.98, 0.27] | -1.82 | *.072* | -0.39 [-0.81, 0.04] | -2.04 [-5.22, 1.14] | -1.27 | .206 | -0.28 [-0.72, 0.16] |
| **PANSS-P** | | | | | | | | |
| OXT | -1.06 [-1.98, -0.14] | -2.3 | **.024** | -0.5 [-0.94, -0.07] | -1.39 [-2.31, -0.48] | -3.02 | **.003** | -0.83 [-1.37, -0.28] |
| PLA | -0.61 [-1.56, 0.33] | -1.29 | .2 | -0.24 [-0.61, 0.13] | -0.57 [-1.54, 0.39] | -1.19 | .238 | -0.28 [-0.76, 0.19] |
| **PANSS-G** | | | | | | | | |
| OXT | -3.12 [-5, -1.24] | -3.3 | **.002** | -0.79 [-1.26, -0.31] | -3.93 [-5.81, -2.05] | -4.16 | **<.001** | -0.89 [-1.32, -0.46] |
| PLA | -1.91 [-3.84, 0.03] | -1.96 | .054 | -0.37 [-0.75, 0.01] | -0.89 [-2.86, 1.08] | -0.9 | .371 | -0.17 [-0.54, 0.2] |
| **CDSS** | | | | | | | | |
| OXT | -1.03 [-2.63, 0.57] | -1.28 | .205 | -0.33 [-0.84, 0.18] | -1.12 [-2.72, 0.48] | -1.4 | .166 | -0.24 [-0.57, 0.1] |
| PLA | -1.08 [-2.77, 0.61] | -1.27 | .207 | -0.4 [-1.03, 0.23] | -0.34 [-2.04, 1.35] | -0.41 | .686 | -0.09 [-0.55, 0.36] |
| **SMQ** | | | | | | | | |
| OXT | 3.11 [-0.88, 7.1] | 1.55 | .124 | 0.38 [-0.11, 0.86] | 0.78 [-3.21, 4.77] | 0.39 | .699 | 0.11 [-0.44, 0.65] |
| PLA | 0.31 [-3.89, 4.5] | 0.15 | .884 | 0.03 [-0.36, 0.42] | -1.15 [-5.35, 3.05] | -0.55 | .587 | -0.09 [-0.43, 0.24] |
| **SMQ Mindful Observation** | | | | | | | | |
| OXT | 1.05 [-0.67, 2.77] | 1.21 | .229 | 0.29 [-0.19, 0.77] | 0.62 [-1.1, 2.34] | 0.72 | .476 | 0.13 [-0.22, 0.47] |
| PLA | -0.12 [-1.95, 1.71] | -0.13 | .899 | -0.03 [-0.46, 0.41] | -0.33 [-2.16, 1.5] | -0.36 | .717 | -0.07 [-0.48, 0.33] |
| **SMQ Letting Go** | | | | | | | | |
| OXT | 0.83 [-0.46, 2.11] | 1.28 | .203 | 0.33 [-0.18, 0.85] | -0.08 [-1.36, 1.2] | -0.12 | .902 | -0.03 [-0.5, 0.44] |
| PLA | 0.27 [-1.08, 1.62] | 0.4 | .688 | 0.08 [-0.31, 0.46] | 0.16 [-1.18, 1.51] | 0.24 | .81 | 0.05 [-0.35, 0.44] |
| **SMQ Absence of Aversion** | | | | | | | | |

| | | | | | | | | |
|---|---|---|---|---|---|---|---|---|
| OXT | -0.42 [-2.29, 1.45] | -0.45 | .654 | -0.08 [-0.44, 0.28] | 0.1 [-1.77, 1.97] | 0.11 | .915 | 0.03 [-0.44, 0.49] |
| PLA | 0.28 [-1.7, 2.27] | 0.29 | .776 | 0.09 [-0.55, 0.73] | -0.04 [-2.02, 1.95] | -0.04 | .97 | -0.01 [-0.41, 0.4] |
| **SMQ Non-judgement** | | | | | | | | |
| OXT | 1.74 [0.17, 3.31] | 2.21 | **.03** | 0.42 [0.04, 0.8] | 0.22 [-1.35, 1.79] | 0.28 | .782 | 0.07 [-0.4, 0.53] |
| PLA | -0.34 [-2, 1.31] | -0.41 | .682 | -0.09 [-0.51, 0.34] | -1.09 [-2.75, 0.57] | -1.31 | .194 | -0.29 [-0.72, 0.15] |
| **PANSS-Marder** | | | | | | | | |
| OXT | -2.08 [-3.29, -0.88] | -3.44 | **.001** | -0.75 [-1.19, -0.32] | -2.46 [-3.67, -1.26] | -4.07 | **<.001** | -0.77 [-1.15, -0.39] |
| PLA | -0.93 [-2.17, 0.31] | -1.49 | .139 | -0.27 [-0.63, 0.09] | -0.73 [-2, 0.53] | -1.15 | .252 | -0.29 [-0.78, 0.21] |
| **PANSS-Wallwork** | | | | | | | | |
| MBGT+OXT | -1.89 [-2.91, -0.87] | -3.69 | **<.001** | -0.85 [-1.3, -0.39] | -2.22 [-3.24, -1.2] | -4.34 | **<.001** | -0.8 [-1.16, -0.43] |
| MBGT+PLA | -0.57 [-1.61, 0.48] | -1.08 | .285 | -0.19 [-0.55, 0.17] | -0.62 [-1.69, 0.45] | -1.16 | .251 | -0.31 [-0.85, 0.23] |

*Note.* MBGT: Mindfulness-based group therapy; OXT: Oxytocin; PLA: Placebo; PANSS-N: Positive and Negative Syndrome Scale – Negative Subscale; BNSS: Brief Negative Symptom Scale; SNS: Self-Evaluation of Negative Symptoms; PANSS-T: Positive and Negative Syndrome Scale – Total Score; PANSS-P: Positive and Negative Syndrome Scale – Positive Subscale; PANSS-G: Positive and Negative Syndrome Scale – General Psychopathology Subscale; CDSS: Calgary Depression Scale for Schizophrenia; SMQ: Southampton Mindfulness Questionnaire. *p-values* below .05 are displayed in bold script, while those below .10 are displayed in italic script.

**Table S3** – *Between Group Changes across Baseline (T0), Post-Intervention (T1), and Four-Week Follow-Up (T2) Including "Living With Partner" as Covariate*

| | **Between Group Effects** | | | | | | | |
|---|---|---|---|---|---|---|---|---|
| | **T1** | | | | **T2** | | | |
| | **Δ [95% CI]** | **t** | **p** | **d [95% CI]** | **Δ [95% CI]** | **t** | **p** | **d [95% CI]** |
| **PANSS-N** | | | | | | | | |
| | -1.61 [-3.65, 0.43] | -1.59 | .119 | -0.28 [-0.64, 0.08] | -1.96 [-4.01, 0.09] | -1.92 | *.061* | -0.37 [-0.76, 0.02] |
| **BNSS** | | | | | | | | |
| | -2.04 [-5.75, 1.68] | -1.1 | .276 | -0.15 [-0.44, 0.13] | -4.54 [-8.26, -0.83] | -2.47 | **.018** | -0.34 [-0.62, -0.06] |
| **BNSS-Anhedonia** | | | | | | | | |
| | -0.33 [-1.87, 1.21] | -0.43 | .667 | -0.09 [-0.52, 0.34] | -1.26 [-2.8, 0.27] | -1.65 | .105 | -0.3 [-0.66, 0.06] |
| **BNSS-Distress** | | | | | | | | |
| | 0.35 [-0.18, 0.87] | 1.32 | .192 | 0.27 [-0.14, 0.68] | 0.09 [-0.43, 0.62] | 0.36 | .724 | 0.07 [-0.31, 0.45] |
| **BNSS-Asociality** | | | | | | | | |
| | -0.4 [-1.32, 0.53] | -0.86 | .397 | -0.17 [-0.57, 0.23] | -0.92 [-1.85, 0.01] | -1.98 | *.053* | -0.36 [-0.72, 0.01] |
| **BNSS Avolition** | | | | | | | | |
| | 0.09 [-0.69, 0.88] | 0.24 | .808 | 0.04 [-0.3, 0.38] | -0.51 [-1.29, 0.27] | -1.32 | .194 | -0.23 [-0.59, 0.12] |
| **BNSS Blunted affect** | | | | | | | | |
| | -1.08 [-2.18, 0.02] | -1.96 | .055 | -0.27 [-0.55, 0.01] | -1.28 [-2.38, -0.18] | -2.34 | **.023** | -0.35 [-0.66, -0.05] |
| **BNSS Alogia** | | | | | | | | |
| | -0.65 [-1.65, 0.35] | -1.31 | .196 | -0.21 [-0.53, 0.11] | -0.68 [-1.68, 0.31] | -1.38 | .175 | -0.21 [-0.52, 0.1] |
| **SNS** | | | | | | | | |
| | 0.53 [-3.02, 4.08] | 0.3 | .766 | 0.07 [-0.43, 0.58] | -1.35 [-4.94, 2.25] | -0.75 | .457 | -0.18 [-0.67, 0.3] |
| **SNS Social withdrawal** | | | | | | | | |
| | 0.21 [-0.72, 1.14] | 0.45 | .653 | 0.13 [-0.45, 0.71] | 0.35 [-0.59, 1.29] | 0.75 | .457 | 0.19 [-0.32, 0.7] |
| **SNS Diminished emotional range** | | | | | | | | |
| | 0.31 [-0.76, 1.37] | 0.57 | .57 | 0.18 [-0.46, 0.82] | -0.49 [-1.57, 0.59] | -0.91 | .365 | -0.24 [-0.76, 0.28] |

| | | | | | | | | |
|---|---|---|---|---|---|---|---|---|
| **SNS Avolition** | | | | | | | | |
| | -0.6 [-1.65, 0.44] | -1.15 | .253 | -0.26 [-0.71, 0.19] | -0.78 [-1.84, 0.28] | -1.46 | .148 | -0.34 [-0.81, 0.12] |
| **SNS Anhedonia** | | | | | | | | |
| | 0.06 [-0.94, 1.07] | 0.12 | .902 | 0.03 [-0.47, 0.53] | -0.33 [-1.35, 0.69] | -0.65 | .518 | -0.15 [-0.61, 0.31] |
| **SNS Alogia** | | | | | | | | |
| | 0.19 [-0.92, 1.3] | 0.34 | .735 | 0.1 [-0.49, 0.7] | -0.44 [-1.56, 0.68] | -0.79 | .434 | -0.23 [-0.8, 0.35] |
| **PANSS-T** | | | | | | | | |
| | -3.28 [-8.49, 1.92] | -1.27 | .211 | -0.24 [-0.63, 0.14] | -6.03 [-11.28, -0.78] | -2.31 | **.025** | -0.49 [-0.91, -0.06] |
| **PANSS-P** | | | | | | | | |
| | -0.45 [-1.84, 0.94] | -0.65 | .52 | -0.09 [-0.38, 0.19] | -0.83 [-2.24, 0.57] | -1.19 | .24 | -0.2 [-0.53, 0.14] |
| **PANSS-G** | | | | | | | | |
| | -0.69 [-3.7, 2.31] | -0.47 | .644 | -0.1 [-0.52, 0.33] | -2.64 [-5.67, 0.39] | -1.75 | *.086* | -0.41 [-0.89, 0.06] |
| **CDSS** | | | | | | | | |
| | 0.12 [-2.25, 2.48] | 0.1 | .922 | 0.03 [-0.55, 0.61] | -0.72 [-3.08, 1.65] | -0.61 | .547 | -0.14 [-0.6, 0.32] |
| **SMQ** | | | | | | | | |
| | 3.15 [-3.15, 9.45] | 1 | .32 | 0.24 [-0.24, 0.71] | 2.25 [-4.06, 8.57] | 0.72 | .477 | 0.15 [-0.26, 0.56] |
| **SMQ Mindful Observation** | | | | | | | | |
| | 0.19 [-2.28, 2.66] | 0.15 | .879 | 0.05 [-0.59, 0.69] | -0.06 [-2.53, 2.41] | -0.05 | .961 | -0.01 [-0.58, 0.55] |
| **SMQ Letting go** | | | | | | | | |
| | 0.69 [-1.21, 2.58] | 0.72 | .472 | 0.17 [-0.3, 0.63] | -0.12 [-2.01, 1.78] | -0.13 | .901 | -0.03 [-0.44, 0.39] |
| **SMQ Absence of aversion** | | | | | | | | |
| | -0.71 [-3.25, 1.84] | -0.55 | .582 | -0.16 [-0.76, 0.43] | 0.14 [-2.41, 2.7] | 0.11 | .91 | 0.03 [-0.51, 0.57] |
| **SMQ Non-judgement** | | | | | | | | |
| | 1.95 [-0.53, 4.43] | 1.58 | .12 | 0.43 [-0.12, 0.98] | 1.2 [-1.29, 3.7] | 0.97 | .336 | 0.23 [-0.25, 0.71] |

| | | | | | | | | |
|---|---|---|---|---|---|---|---|---|
| **PANSS-Marder** | | | | | | | | |
| | -0.7 [-2.58, 1.18] | -0.75 | .459 | -0.13 [-0.49, 0.22] | -1.32 [-3.22, 0.58] | -1.4 | .169 | -0.23 [-0.57, 0.1] |
| **PANSS-Wallwork** | | | | | | | | |
| | -0.87 [-2.42, 0.67] | -1.14 | .261 | -0.18 [-0.5, 0.14] | -1.16 [-2.72, 0.4] | -1.49 | .142 | -0.24 [-0.55, 0.08] |

*Note.* MBGT: Mindfulness-based group therapy; OXT: Oxytocin; PLA: Placebo; PANSS-N: Positive and Negative Syndrome Scale – Negative Subscale; BNSS: Brief Negative Symptom Scale; SNS: Self-Evaluation of Negative Symptoms; PANSS-T: Positive and Negative Syndrome Scale – Total Score; PANSS-P: Positive and Negative Syndrome Scale – Positive Subscale; PANSS-G: Positive and Negative Syndrome Scale – General Psychopathology Subscale; CDSS: Calgary Depression Scale for Schizophrenia; SMQ: Southampton Mindfulness Questionnaire. *p-values* below .05 are displayed in bold script, while those below .10 are displayed in italic script.
